# Enacting national social distancing policies corresponds with dramatic reduction in COVID19 infection rates

**DOI:** 10.1101/2020.04.23.20077271

**Authors:** Daniel J. McGrail, Jianli Dai, Kathleen M. McAndrews, Raghu Kalluri

## Abstract

The outbreak the SARS-CoV-2 (CoV-2) virus has resulted in over 2.5 million cases of COVID19, greatly stressing global healthcare infrastructure. Lacking medical prophylactic measures to combat disease spread, many nations have adopted social distancing policies in order to mitigate transmission of CoV-2. While mathematical models have suggested the efficacy of social distancing to curb the spread of CoV-2, there is a lack of systematic studies to quantify the real-world efficacy of these approaches. Here, we quantify the spread rate of COVID19 before and after national social distancing measures were implemented in 26 nations and compare this to the changes in COVID19 spread rate over equivalent time periods in 27 nations that did not enact social distancing policies. We find that social distancing policies significantly reduced the COVID19 spread rate. Using mixed linear regression models we estimate that social distancing policies reduced the spread of COVID19 by 66%. These data suggest that social distancing policies may be a powerful tool to prevent spread of COVID19 in real-world scenarios.

## INTRODUCTION

COVID19, caused by the novel coronavirus SARS-CoV-2 (CoV-2) (Zhou et al., 2020), was declared a global pandemic by the World Health Organization on March 11^th^ 2020. As of April 21^st^ 2020, the disease had spread to over 2.5 million cases worldwide, straining the global healthcare infrastructure, and is rapidly becoming a leading cause of death (Keating and Esteban, 2020). Medical interventions to address COVID19 are lacking both in the context of prophylactic vaccination approaches, as well as pharmaceutical interventions to treat infected patients. Some countries such as Singapore have demonstrated the feasibility of contact tracing when sufficient testing capacities are available to quarantine exposed individuals and contain disease spread (Anderson et al., 2020). In nations where contact tracing is not feasible, “social distancing” policies have been implemented by a subset of governments. These policies include closure of non-essential workplaces and schools, as well as policies on physical spacing when in public aimed at preventing viral transmission to reduce the number of new COVID19 cases.

Mathematical models of COVID19 spread have indicated that social distancing policy will likely reduce the spread of CoV-2 (Kissler et al., 2020). Case reports from regions nearby Hubei province where the virus originated have demonstrated social distancing policies will reduce viral transmission (Leung et al., 2020), however the efficacy of the approach has yet to be evaluated on a global scale. Here, using data from 53 nations we assess the efficacy of social distancing approaches and find that social distancing significantly prevents spread of COVID19, resulting in an estimated 66% reduction in new COVID19 cases.

## METHODS

### Data Sources

All data were acquired on April 21^st^ 2020. Daily case numbers for COVID19 and population numbers were acquired from https://www.worldometers.info/coronavirus/. Social distancing policies were acquired from https://auravision.ai/covid19-lockdown-tracker/. Countries were considered for inclusion if they had over 500 cases of COVID19, and at least 10 days of data prior to and following implementation of social distancing policy (or equivalent time period in countries without a social distancing policy). Countries with regional social distancing policies were excluded from analysis.

### Data Analysis

All data analysis was performed in MATLAB R2019a. For per country COVID19 spread rates, data were modeled to an exponential growth equation *Cases* ∼ *Cases0*·exp(*k*_spread_·*t*), where Cases is cases per million inhabitants, Cases0 is the number of cases per million inhabitants at the initial time point, k_spread_ is the COVID19 spread rate, and t is the time point in days. The initial time point was considered as when countries exceeded 1 case per million inhabitants. Post social distancing was considered 7 days after social distancing policy was enacted. For global modeling of COVID19 spread, the above equation was utilized is a generalized linear mixed-effects model taking each country as a random effect. To estimate prevented number of cases, the change in COVID19 spread rate in countries with social distancing policies was equated to that of those without social distancing. Final total number cases was determined after 14 days of spread.

## RESULTS

In order to assess the efficacy of social distancing policies, we began by quantifying the COVID19 spread rate as defined by an exponential growth function both before and after social distancing policies were enacted. As the rate of disease spreading may be influenced by other temporal features such as weather patterns or disproportional infection of immunocompromised individuals at early stages, we further quantified COVID19 spread rates over time-matched intervals for nations without social distancing policies. This approach is illustrated by comparison of Norway and Sweden, two countries with similar demographics and healthcare policies (**Figure 1A-D**). Norway enacted social distancing policies on March 12^th^ 2020, whereas no such policies were enacted in Sweden. In Norway, pre-social distancing COVID19 spread rate was 0.28 (**Figure 1A**), dropping to 0.07 following social distancing (**Figure 1B**) for a net change of -0.21. When analyzing spread rate in Sweden over equivalent time periods we found that Sweden only dropped from 0.13 (**Figure 1C**), to 0.10 (**Figure 1D**) for a net change of -0.03, 7-fold lower than observed in Norway. Normalization of spreading rates on a per country basis helps to mitigate variances based on population demographics, climate, healthcare infrastructure, and other factors that vary across nations. We applied this approach to 53 countries total for which sufficient data was available (**Figure 1E**). Comparing the resulting changes in COVID19 spread rate we found that countries which enacted social distancing policies demonstrated a significantly larger reduction in COVID19 spread rates (p = 5.3×10^−4^, **Figure 1F**).

**Figure 1.**
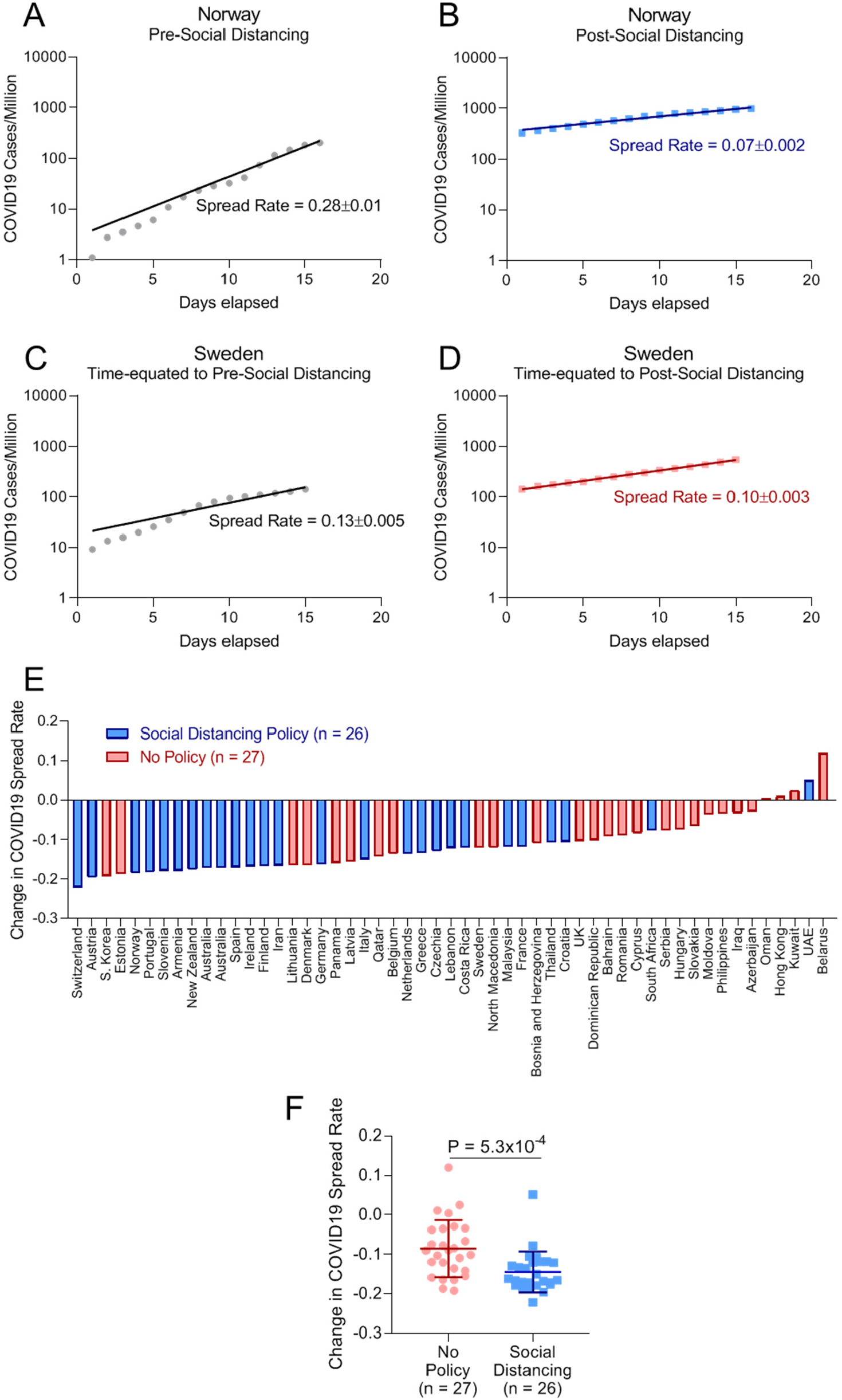
Reduction in COVID19 spread rates in countries with social distancing. **(A-D)** Fitting of COVID19 spread rates in Norway (**A-B**) prior to social distancing policy implementation (**A**) and after social distancing policy implementation (**B**) as well as Sweden (**C-D**) in a time period equated to pre-social distancing in Norway (**C**) and post-social distancing in Norway (**D**). (**E**).Change in COVID19 spread before and after social distancing in countries with social distancing policies (blue), or time equated periods in countries with no social distancing policies (red). (**F**).Dot plot of values shown in (**E**) showing reduction in COVID19 spread following implementation of social distancing policies. Rank-sum test.

In order to assess the benefit of this reduction in COVID19 spread rates, we refined our calculations using generalized linear mixed effect models taking each country as a random effect. This approach again recapitulated the larger drop in COVID19 spread rates observed in countries with social distancing policies (**Figure 2A**). To quantify the healthcare benefits resulting from this reduction in spread rate, we calculated COVID19 spread rates in countries with social distancing policies both using the model directly fit to the data, as well as correcting for the reduction in COVID19 spread rate observed following implementation of social distancing policies (**Figure 2B**). After accounting for number of inhabitants, this approach indicates that over a two week period social distancing prevented over half a million cases of COVID19 in the 26 nations with social distancing policies, representing a 66% reduction in new COVID19 cases (**Figure 2C**).

**Figure 2.**
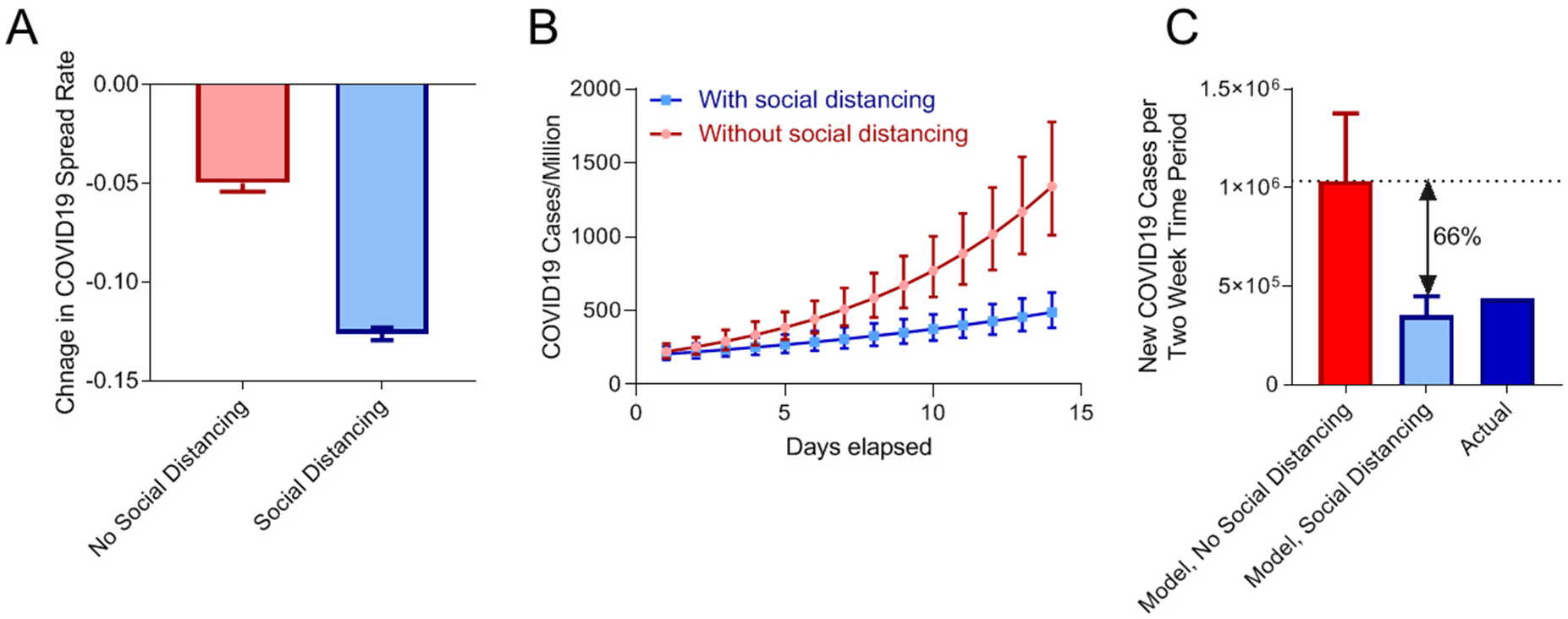
Prevention of new COVID19 cases by social distancing policies. (**A**).Change in COVID19 spread rates following implementation of social distancing policies (blue), or time-equated periods in countries with no social distancing policies (red). Data were fit using a generalized linear mixed-effects model taking each country as a random effect. Bars represent change in regression coefficient ± standard error. (**B**).Modeling of new COVID19 cases per million inhabitants in countries with social distancing policies using either model fit directly to data following implementation of social distancing policies (blue), or after correcting for the observed reduction in COVID19 spread rates associated with social distancing policies (red). Points represent model COVID19 cases per million ± standard error. (**C**).Estimation of total new cases over a two-week period if countries had not implemented social distancing policies (red), with implementation of social distancing policies (light blue), and actual new cases (dark blue). Bars represent number of COVID19 cases in the countries with social distancing policies ± standard error.

## DISCUSSION

Our analysis of COVID19 spread rates in 53 countries quantifies the effects of social distancing at preventing CoV-2 transmission. While numerous factors such as population density, healthcare infrastructure, testing rates, climate, population characteristics, and more, likely contribute to rate of COVID19 spread, this study focused on the change in COVID19 spread rate following implementation of social distancing policies as an internal control for these variables. This analysis found that social distancing policies significantly reduced spreading rates compared to time-matched intervals in nations that did not implement social distancing policies. While social distancing policies may have negative economic impacts (Eichenbaum et al., 2020), this analysis suggests that this containment approach has yielded significant positive health outcomes.

As testing capacities increase, contact tracing may provide an alternative to minimize economic impacts by only quarantining those who are exposed to infected individuals. This approach has already demonstrated high efficacy in the context of nations with high testing capacity (Anderson et al., 2020). Technological advancements, such as potential phone-based contact tracing Apps, may further enhance the benefit achieved by contact tracing approaches (Ferretti et al., 2020).

The emergence of further refined data will allow for more precise quantification of various approaches to mitigate the spread of COVID19. This analysis based on 53 countries indicates that social distancing can serve as a critical preventative measure until prophylactic measures and larger population immunity are generated.

## Data Availability

All information is publicly available.

https://www.worldometers.info/coronavirus/

